# Exploring the genetic overlap of suicide-related behaviors and substance use disorders

**DOI:** 10.1101/2021.04.05.21254944

**Authors:** Sarah M.C. Colbert, Alexander S. Hatoum, Andrey Shabalin, Hilary Coon, Elliot C. Nelson, Arpana Agrawal, Anna R. Docherty, Emma C. Johnson

## Abstract

**Background:** Suicide-related behaviors are heterogeneous and transdiagnostic, and may demonstrate varying levels of genetic overlap with different substance use disorders (SUDs).

**Methods:** We used linkage disequilibrium score regression, genomic structural equation models, and Mendelian Randomization to examine the genetic relationships between several SUDs and suicide-related behaviors. Our analyses incorporated summary statistics from the largest genome-wide association studies (GWAS) of problematic alcohol use (PAU), the Fagerström Test for Nicotine Dependence (FTND), cannabis use disorder (CUD), and opioid use disorder (OUD; Ns ranging from 46,213-435,563) and GWAS of ever self-harmed, suicide attempt, and suicide death (Ns ranging from 18,223-117,733). We also accounted for genetic liability to depression (N=500,199) and risk tolerance (N=315,894).

**Results:** Suicide-related behaviors were significantly genetically correlated with each other and each SUD, but there was little evidence of causal relationships between the traits. Simultaneously correlating a common SUD factor with each specific suicide indicator while controlling for depression and risk tolerance revealed significant, positive genetic correlations between the SUD factor and suicide-related behaviors (r_g_ = 0.26-0.45, se=0.08-0.09). In the model, depression’s association with suicide death (β = 0.42, se = 0.06) was weaker compared to ever-self harmed and suicide attempt (β = 0.58, se=0.05 and β = 0.50, se=0.06, respectively).

**Discussion:** We identify a general level of genetic overlap between SUDs and suicide-related behaviors which is independent of depression and risk tolerance. Additionally, our findings suggest that genetic and behavioral contributions to suicide death may somewhat differ from non-lethal suicide-related behaviors.

## Introduction

Self-harming behaviors, encompassing suicidal and non-suicidal acts, are components of various psychiatric syndromes. Suicide was the second leading cause of death in individuals aged 10-34 years in 2018 (Xu et al., 2020). Furthermore, nearly 1.5 million adults reported a suicide attempt in 2018, with 443,000 adults requiring overnight hospitalization for their attempt (Centers for Disease Control and Prevention, 2020; SAMHSA, 2019). Substance use problems often precede and co-occur with suicidal thoughts and behaviors (Poorolajal et al., 2016). For example, self-reported problematic alcohol use was associated with a 5-fold increase in risk for suicide attempt and growing up with a problem drinker or drug user in the immediate family doubled the odds for suicide attempt (Dube et al., 2001).

Both suicide-related behaviors and substance use disorders (SUDs) are heritable (h^2^ = 17 – 55% and 39 – 72%, respectively; (Ducci & Goldman, 2012; Ruderfer et al., 2020; Statham et al., 1998)), polygenic traits, with evidence of genetic overlap between them from both traditional twin and family studies as well as more recent genome-wide association studies (GWAS). For example, twin studies have demonstrated evidence of shared genetic underpinnings for suicide-related behaviors and cannabis dependence (Lynskey et al., 2004), alcohol use disorder (Edwards et al., 2020) and tobacco smoking (Evins et al., 2017) and family studies have shown that substance-dependent individuals who attempt suicide are more likely to have first-degree relatives with a history of suicide-related behaviors (Brent & Mann, 2005). Several GWAS of suicide-related behaviors have also documented significant genetic overlap with alcohol use phenotypes; for example, Docherty et al. (2020) found that polygenic scores of suicide death were significantly associated with greater alcohol use, and Mullins et al. (preprint; 2020) found that suicide attempt was positively genetically correlated with both alcohol dependence and the Problems subscale of the Alcohol Use Disorders Identification Test (AUDIT-P). While there is substantial evidence of genetic sharing amongst suicide-related behaviors and SUDs, mechanisms underlying this overlap are yet to be identified. Through observational and longitudinal studies (Berlin et al., 2015; Flensborg-Madsen et al., 2009; Gobbi et al., 2019; Malone et al., 2003), researchers have hypothesized that causality may be underlying the relationships between suicide-related behaviors and SUDs. Only recently have genetic studies been able to provide further support for this hypothesis, with a recent Mendelian randomization study suggesting possible causal roles of cannabis, alcohol and smoking on suicide attempt (Orri et al., 2020).

Heterogeneity amongst SUDs and their distinct genetic relationships with other dimensions of mental health are becoming increasingly better understood (Abdellaoui et al., 2021; Hatoum et al., 2021). Recently, Hatoum et al. (preprint; 2021) proposed a unidimensional factor for characterizing substance use disorders, specifically problematic alcohol use (PAU), cannabis use disorder (CUD), opioid use disorder (OUD) and problematic tobacco use. Notably, the authors found that the substance use disorder factor was correlated with non-substance use psychopathology, but retained significant independent variance.

In contrast, the genetic architectures of suicide-related behaviors are less well-studied. Some have tested whether suicide attempt and suicide death represent a continuum of genetic vulnerability; the evidence thus far does not support this hypothesis (Kendler et al., 2020). Still, twin studies have suggested some genetic commonality between suicidal ideation and attempt, as well as with non-suicidal self-harm (MacIejewski et al., 2014; Richmond-Rakerd et al., 2019). Only recently have GWAS begun to directly compare suicide-related behaviors, with results indicating both shared and unique genetic components amongst suicide-related behaviors, as indexed by their genetic relationships with other forms of psychopathology (Campos et al., 2020; Strawbridge et al., 2019). For example, Strawbridge et. al (2019) observed stronger genetic correlations between self-harm and major depressive disorder (MDD) than between suicidal ideation or attempt and MDD. However, self-harm and suicidal ideation or attempt also showed similar genetic correlations with other psychiatric disorders such as schizophrenia (Strawbridge et al., 2019). Most of these suicide related GWAS examine genetic overlap with a variety of psychiatric traits, yet genetic overlap with substance use behaviors (other than alcohol use) is largely unreported.

Large scale GWAS of both SUDs and suicide-related behaviors now enable an examination of the genetic architecture underlying their comorbidity. An important consideration in such analyses is the role of MDD – suicidal thoughts and behaviors are among diagnostic criteria for MDD, even though they are pervasive in other psychiatric syndromes (American Psychiatric Association, 2013). Among psychiatric disorders, depression shares one of the highest genetic correlations with both suicide attempt (Mullins et al., 2020; Ruderfer et al., 2020) and SUDs (Abdellaoui et al., 2021), yet whether the genetic relationship between SUDs and suicide-related behaviors extends beyond depression remains understudied. For instance, Mullins et al. (preprint; 2020) estimated genetic correlations with suicide attempt before and after conditioning suicide attempt on MDD. They found that removing the genetic overlap with MDD generally weakened the genetic correlations between suicide attempt and psychiatric disorders by 33% on average, but did not significantly affect the genetic correlations between suicide attempt and non-psychiatric phenotypes (including alcohol use and smoking). However, in another study, alcohol use and suicide attempt were found only to be related through their common associations with depression (Grazioli et al., 2018). In addition to MDD, risk-taking behaviors are also correlated with both SUDs (Johnson et al., 2020; Zhou, Sealock, et al., 2020) and suicide attempt (Mullins et al., 2020), and could be contributing to observed genetic correlations among these traits. Deeper phenotyping of lethality of suicide attempts have revealed a role for impulsivity, albeit mixed (Baca-Garcia et al., 2005) and the role of risk-taking as a genetic co-contributor to SUD and suicide remains unknown. Including depression and risk tolerance as genetic covariates provides opportunities to examine their influence on this comorbidity and whether the genetic overlap between SUD and suicide-related behaviors is partly independent of these shared traits.

We examined the genetic relationships between SUDs and suicide-related behaviors by (i) estimating pair-wise genetic correlations amongst four SUD phenotypes (opioid use disorder (OUD), cannabis use disorder (CUD), nicotine dependence [estimated using the Fagerström Test for Nicotine Dependence; FTND] and problematic alcohol use (PAU)), three suicide-related behaviors (suicide death, suicide attempt and ever self-harmed) and two additional phenotypes (depression and risk tolerance) that may be relevant to these associations; (ii) testing for a causal influence of SUDs on suicide-related behaviors with causal analysis using summary effect estimates (CAUSE; Morrison et al., 2020); and (iii) applying genomic structural equation models (gSEM; Grotzinger et al., 2019) to the genetic correlation matrix of these traits to identify the genetic factor structure.

## Methods

### Samples

We used publicly available summary statistics from GWA studies of four SUDs and three suicide-related behaviors (Table 1). Summary statistics for the SUDs came from four studies, which to our knowledge are the largest available GWAS for each phenotype. We chose to use these phenotypes specifically as their contributions to a common genetic addictions factor are well-studied (Hatoum et al., 2021).

**Table 1.**
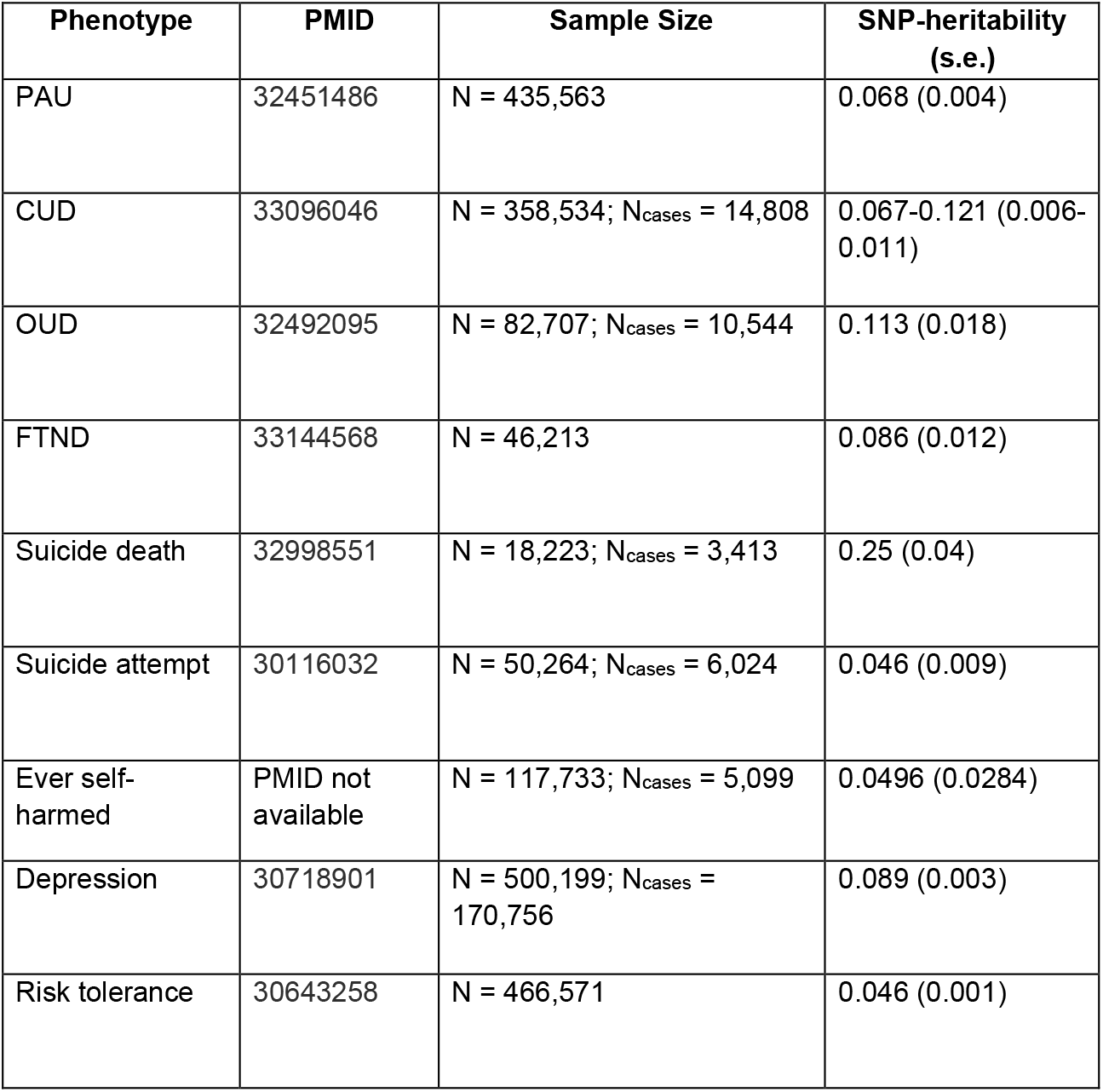
Descriptive statistics for suicide-related behaviors, substance use disorders, and related risk factor (depression, risk tolerance) genome-wide association studies. Ever self-harmed summary statistics came from the publicly-available Neale Lab GWAS (http://www.nealelab.is/uk-biobank) but have not been published and thus have no PMID available.

- *Problematic alcohol use:* We used summary statistics for PAU which came from a GWAS meta-analysis of data from the Psychiatric Genomics Consortium (PGC), Million Veteran Program (MVP) and UK Biobank (N=435,563) (Zhou, Sealock, et al., 2020).
- *Cannabis use disorder:* Summary statistics for CUD (N=358,534) were derived from a meta-analysis of GWAS data from the PGC, the Lundbeck Foundation Initiative for Integrative Psychiatric Research (iPSYCH) and deCODE Genetics (Johnson et al., 2020).
- *Opioid use disorder:* OUD summary statistics (N=82,707) came from a GWAS meta-analysis of OUD in data from MVP, Yale-Penn and Study of Addiction: Genetics and Environment (Zhou, Rentsch, et al., 2020).
- *Nicotine dependence:* Nicotine dependence summary statistics (N=46,213) came from a GWAS of the Fagerström Test for Nicotine Dependence (FTND) from the Nicotine Dependence GenOmics (iNDiGO) Consortium (Quach et al., 2020).

We used the phenotypes ever self-harmed, suicide attempt and suicide death from three independent studies to capture the variation in suicide-related behavior (Table 1).

- *Ever self-harmed*: Summary statistics (N=117,733) were derived from a GWAS (conducted by the Neale lab: http://www.nealelab.is/uk-biobank) of field 20480 in the UK Biobank (*Imputed Genotypes from HRC plus UK10K & 1000 Genomes Reference Panels as Released by UK Biobank in March 2018*., 2018) which is a self-report measure of whether an individual has ever self-harmed, with or without the intent of ending one’s life. Self-harm can include a range of behaviors, from self-cutting and hitting to ingesting medication, alcohol or illicit drugs in excess to endanger oneself (UKB Data-Field 20553). Due to the ambiguity of intent in this measure, cases include individuals who may have attempted suicide.
- *Suicide attempt*: Summary statistics (N=50,264) came from a GWAS of diagnoses of suicide attempt using International Classification of Diseases (ICD-10) codes from iPSYCH data (Erlangsen et al., 2020). In addition to ICD-10 codes for suicide attempt (ICD-10:X60-X84), proxies for suicide attempt included individuals where the “reason for contact”-variable indicated suicide attempt and individuals with a main diagnosis of a mental disorder accompanied by a diagnosis of a self-harm behavior.
- *Suicide death*: Summary statistics for suicide death (N=18,223) were derived from data from the Utah Office of the Medical Examiner (Docherty et al., 2020).

In addition, we included summary statistics for depression and risk tolerance (i.e., willingness to take risks) as covariates in our models as these traits are genetically correlated with both suicide-related behaviors and SUDs. We used depression summary statistics (N=500,199) from a meta-analysis of GWAS data from the UKB and PGC (Howard et al., 2019). Risk tolerance summary statistics (N=466,571) were derived from a meta-analysis of general risk tolerance in the UKB and 10 other cohorts (Karlsson Linnér et al., 2019). The specific measure of this phenotype varied across the cohorts, yet all used similar measures of an individual’s general willingness to take risks. We chose to use this measure of risk-taking over other available options as other risk phenotypes (e.g., the first principal component of four risky behaviors in the UK Biobank; Karlsson Linnér et al., 2019) often include measures relating to substance use, which could potentially inflate the genetic relationship between risk-taking and SUDs in this study.

### Quality Control Procedures

Prior to use in linkage disequilibrium (LD) Score Regression (LDSC) and gSEM, summary statistics were filtered to HapMap3 SNPs, SNPs with INFO score > 0.9 and minor allele frequency < 0.01 using the “munge” function of LDSC (Bulik-Sullivan et al., 2015).

### Genetic correlations

We used LDSC (Bulik-Sullivan et al., 2015) to estimate pairwise genome-wide genetic correlations (r_g_) between SUDs, suicide-related behaviors and covariate (i.e., depression, risk tolerance) phenotypes (Figure 1). We used the Benjamini-Hochberg false discovery rate (FDR) procedure to account for multiple testing (Benjamini & Hochberg, 1995); an FDR < 5% was used to determine significance. We used the jackknife estimate of the difference between genetic correlations (H_0_: r_g_(A,B) - r_g_(A,C) = 0) to determine if two correlations were significantly different from each other (Coleman et al., 2020).

**Figure 1.**
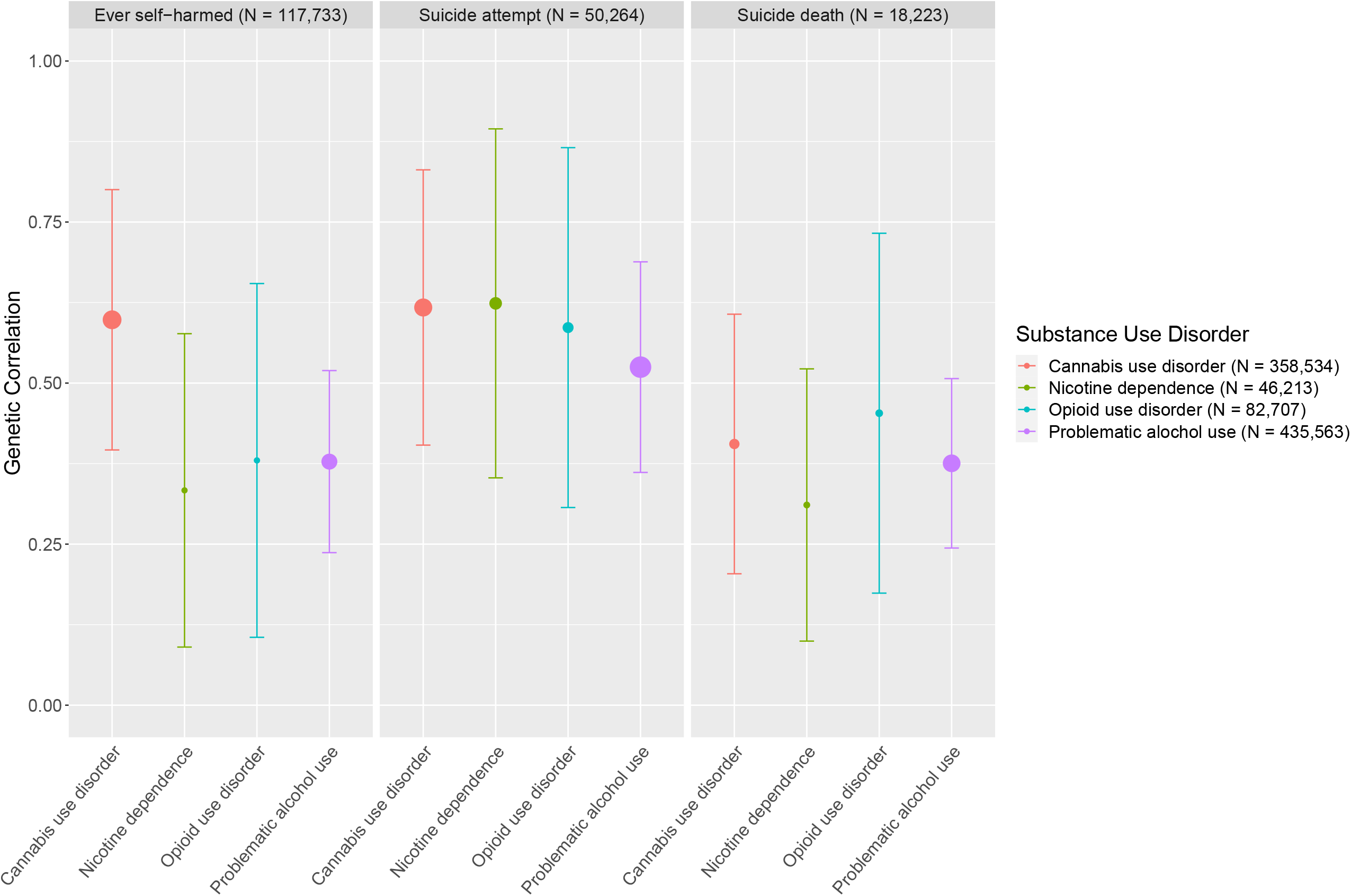
Genetic correlations amongst suicide-related behaviors and SUDs. Error bars represent 95% confidence intervals. All genetic correlation estimates were significant (FDR < 5%). The size of points corresponds to the -log(p-value), with larger points indicating more significant p-values.

### Mendelian Randomization using CAUSE

The causal relationship between SUD and suicide-related behavior phenotypes, in which SUDs are causal for suicide-related behaviors, was tested using CAUSE (Morrison et al., 2020). Causality was tested between each SUD-suicide-related behavior pair. CAUSE, compared to traditional Mendelian randomization methods, is advantageous in that it accounts for correlated horizontal pleiotropic effects and uses a less stringent p-value threshold (p < 1e^-3^) to incorporate data from more variants across the genome. CAUSE constructs two nested models: a sharing model and causal model. Both models allow for horizontal pleiotropic effects; however, only the causal model includes a causal effect parameter. CAUSE compares the sharing and causal models and then computes a z-score that can be compared to a normal distribution. We calculated one-sided p-values corresponding to the z-scores in order to test the null hypothesis that the sharing model fits the data at least as well as the causal model. Significant p-values therefore indicate the presence of a causal effect.

### Genomic structural equation modeling

To estimate the genetic overlap between and within SUDs and suicide-related behaviors and speculate on latent transdiagnostic risk factors, we used gSEM (Grotzinger et al., 2019) to construct and assess the fit of a common factor and correlated two-factor model (Figures S1-S2). We performed a chi-squared difference test to compare model fit. The correlated two-factor model consisted of two latent factors: an SUD factor and a suicide-related behaviors factor (Figure S2). Similar to Hatoum et al., all four SUD phenotypes (PAU, CUD, OUD and FTND) were allowed to load onto the SUD factor while ever self-harmed, suicide attempt and suicide death were allowed to load onto the suicide-related behaviors factor. We also constructed a model in which we correlated the SUD latent factor with each suicide-related behavior individually (Figure S3).

Depression and risk tolerance were added as covariates to the final gSEM (Figure 2). To do so, depression and risk tolerance were made exogenous predictors of all indicators, i.e. all indicators were regressed on depression and risk tolerance and these endogenous variables were simultaneously used for model building. We used equality constraints and chi-squared difference tests to determine whether correlations and associations in the model were significantly different from each other. ln order to achieve proper model identification, we used unit variance identification in all models, constraining the variances of the latent factors to be 1.

**Figure 2.**
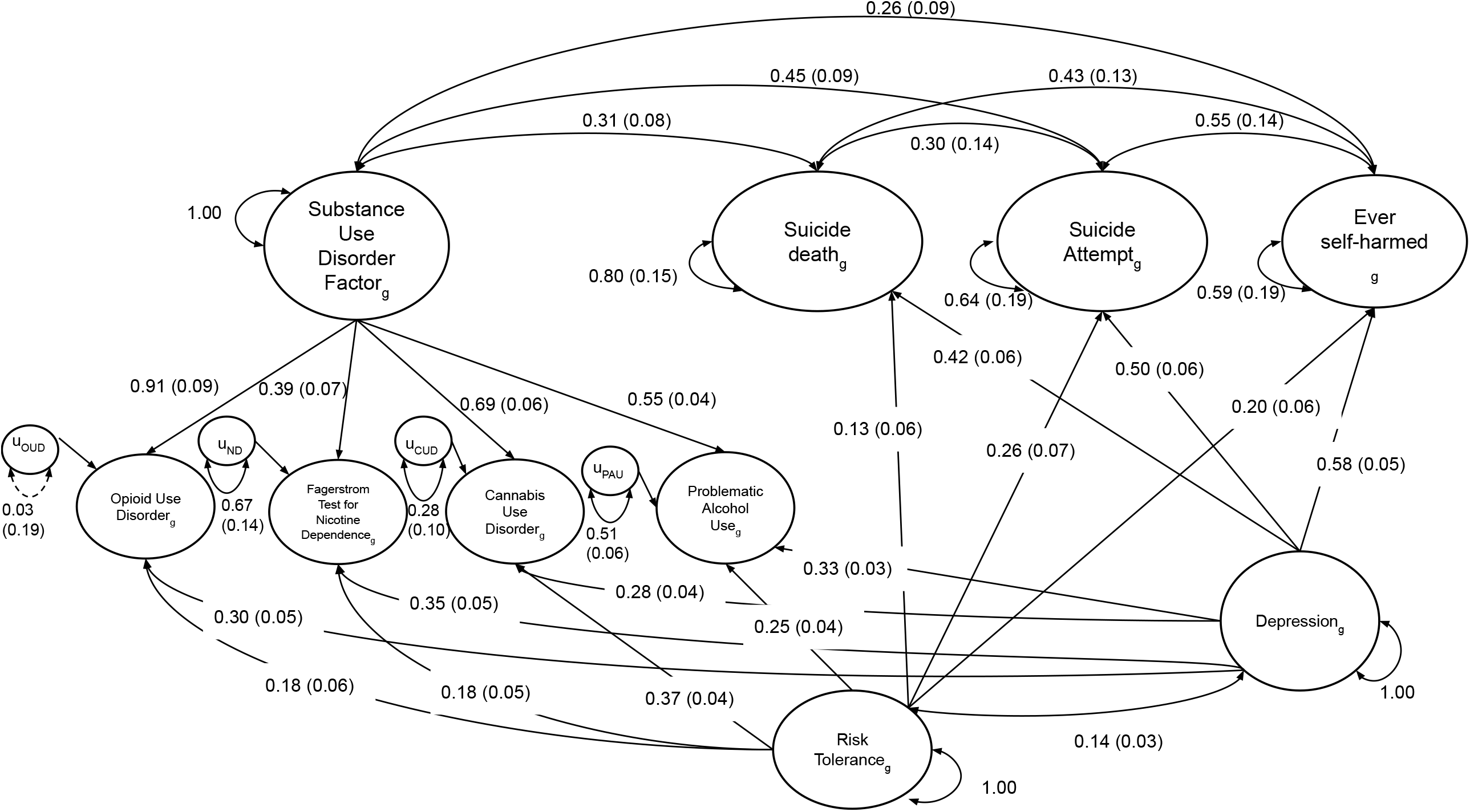
Common SUD and specific suicide-related behaviors model with covariates results. Parameter estimates are standardized with standard errors in parentheses. Solid black lines are used to indicate significance (p < 0.05) while dotted black lines represent non-significant paths. Model fit was excellent (CFI > 0.99 and SRMR < 0.04): χ^2^(11) = 9.16, p_chisq_ = 0.61, AIC = 77.16, CFI = 1.00, SRMR = 0.036.

## Results

### Genetic correlations

As expected, we observed significant positive genetic correlations within the group of SUDs (r_g_ = 0.33-0.79, se = 0.04-0.13, p =1.0×10^−43^-9.3×10^−5^) and within the group of suicide-related behaviors (r_g_ = 0.57-1.05^1^, se = 0.16-0.19, p = 4.1×10^−8^-1.0×10^−4^; Table S1). Among SUDs, PAU, CUD and FTND all correlated the strongest with OUD (r_g_ = 0.50-0.79, se = 0.06-0.13, p = 7.7×10^−29^-9.3×10^−5^), relative to correlations amongst themselves. Suicide attempt and ever self-harmed (which includes suicide attempts) correlated more strongly with each other (r_g_ = 1.05, se = 0.19, p = 4.1×10^−8^) than with suicide death (r_g_ = 0.60, se = 0.16, p = 1.0×10^−4^ and r_g_ = 0.77, se = 0.16, p = 2.1×10^−6^ repectively). Both depression and risk tolerance showed significant genetic correlations with SUDs (r_g_ = 0.22-0.40, se = 0.03-0.05, p = 7.1×10^−38^-4.2×10^−5^) and suicide-related behaviors (r_g_ = 0.20-0.66, se = 0.05-0.08, p = 5.5×10^−18^-4.0×10^−4^); however, depression generally showed stronger genetic correlations compared to risk tolerance (Table S1) for both SUDs and suicide-related phenotypes. Within suicide-related behaviors, depression shared the highest genetic correlation with ever self-harmed (r_g_ = 0.66, se = 0.08, p = 5.5×10^−18^) and weakest with suicide death (r_g_ = 0.42, se = 0.05, p = 8.1×10^−17^).

Despite observing significant and positive genetic correlations across all SUDs and suicide-related behaviors (r_g_ = 0.31-0.62, se = 0.07-0.14, p = 7.2×10^−3^-3.2×10^−10^; Figure 1), we did notice some nominal differences. Of the four SUDs, ever self-harmed correlated most strongly with CUD (r_g_ = 0.60, se = 0.10, p = 6.50e^-9^), and suicide death correlated most strongly with OUD (r_g_ = 0.45, se = 0.17, p = 2.1e^-3^; Figure 1, Table S1); however, the suicide-related behaviors GWAS were underpowered to detect significant differences between these correlations.

### Mendelian Randomization using CAUSE

For each SUD-suicide-related behavior pair, results indicated that the sharing model did not fit significantly worse than the causal model (all p > 0.05; Table S2), therefore, we failed to detect a significant causal effect of any SUD on any of the suicide-related behaviors.

### Genomic structural equation modeling

First, we tested a model in which all four SUDs and all three suicide-related behavior phenotypes were allowed to load onto a single common latent factor (Figure S1). Model fit was adequate (χ^2^(14) = 33.99, p_chisq_ = 0.002, Akaike Information Criterion (AIC) = 61.99, Comparative Fit Index (CFI) = 0.967, Standardized Root Mean Square Residual (SRMR) = 0.119), and standardized loadings indicated that OUD and CUD loaded most strongly onto the common factor, while FTND and suicide death loaded the weakest. We then separated the SUD and suicide-related phenotypes into two latent factors, re-specifying a correlated two-factor model (Figure S2). Model fit was good (χ^2^(13) = 13.90, p_chisq_ = 0.38, AIC = 43.90, CFI = 1, SRMR = 0.065). The correlated two-factor model fit significantly better than the common factor model 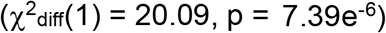. However, the strong loading of suicide attempt (χ = 1.00; Figure S2) in the correlated two-factor model suggested that the suicide-related behaviors factor was isomorphic with suicide attempt (i.e., the genetics of suicide attempt recapitulated the genetic covariance between self-harm, suicide attempt and suicide death). Therefore, instead of imposing a latent structure, we individually correlated each suicide-related behavior with the SUD factor (Figure S3). Model fit was still good (χ^2^(11) = 10.06, p_chisq_ = 0.53, AIC = 44.06, CFI = 1, SRMR = 0.053) and each suicide-related behavior was significantly associated (r_g_ = 0.50-0.72) with the SUD factor.

To examine the genetic relationship between SUDs and suicide-related behaviors unique of their genetic overlap with depression and risk tolerance, we included these two traits as covariates (Figure 2). Model fit improved (χ^2^(11) = 9.16, p_chisq_ = 0.61, AIC = 77.16, CFI = 1, SRMR = 0.036), therefore we chose to move forward with this common SUD and specific-suicide-related behaviors model with covariates (Figure 2).

The genetic relationships between covariates, SUDs, and suicide-related behaviors closely matched the patterns observed in the genetic correlation matrix. Depression was significantly associated with all phenotypes, but showed greater associations with the suicide-related behaviors (β = 0.42-0.58) than with SUD indicators (β = 0.28-0.35), on average. Risk tolerance was also significantly associated with all phenotypes (β = 0.13-0.37). Notably, genetic correlations amongst suicide-related behaviors decreased when covariates were added to the model (r_g_ = 0.30-0.55), as did the genetic correlations between the SUD factor and suicide-related behaviors (r_g_ = 0.26-0.45).

To test whether the SUD factor showed significantly different correlations with the three suicide-related behaviors, we compared our primary model, in which correlations between the SUD factor and individual suicide-related behaviors were estimated freely, to a model which used equality constraints (Figure S4; χ^2^(13) = 13.43, p_chisq_ = 0.41, AIC = 77.43, CFI = 0.998, SRMR = 0.042). A chi-squared difference test was non-significant 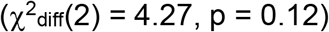, suggesting that genetic correlations with the SUD factor did not significantly differ amongst the specific suicide-related behaviors (r_g_ = 0.34).

Equality constraints were also used to compare the relationships between depression and risk tolerance with individual SUDs and suicide-related phenotypes. Applying equality constraints to the associations between depression and all three-suicide related behaviors (Figure S5) resulted in good model fit (χ^2^(13) = 22.44, p_chisq_ = 0.05, AIC = 86.44, CFI = 0.991, SRMR = 0.043); however, a chi-squared difference test revealed that the model without constraints fit significantly better 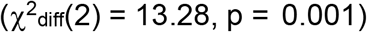, suggesting that the higher genetic correlation between depression and self-harm was statistically meaningful. Similarly, we used equality constraints to compare the associations between risk tolerance and SUDs and found that while model fit was adequate (Figure S6; χ^2^(14) = 49.08, p_chisq_ = 8.71e^-6^, AIC = 111.08, CFI = 0.969, SRMR = 0.047), a chi-squared difference test once again suggested that the model without constraints fit significantly better 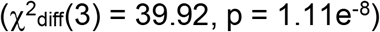 and that risk tolerance was more strongly associated with CUD and PAU than OUD and FTND (Figure S6). The model without constraints was also compared to and fit better 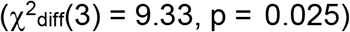 than a model in which the associations between depression and SUDs were constrained to be equal (Figure S7; χ^2^(14) = 18.49, p_chisq_ = 0.19, AIC = 80.49, CFI = 0.9934, SRMR = 0.042). Lastly, we tested equality constraints on the associations between risk tolerance and the suicide-related behaviors (Figure S8; χ^2^(13) = 14.33, p_chisq_ = 0.35, AIC = 78.33, CFI = 0.997, SRMR = 0.039). The non-significant chi-squared difference test 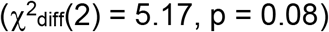 suggested that the constraints did not significantly worsen model fit and therefore the associations between risk tolerance and suicide-related behaviors were not significantly different.

## Discussion

In this study, we used summary statistics from the current largest GWAS of SUDs and suicide-related behaviors to examine the genetic overlap between these psychiatric domains, while accounting for the genetics of depression and risk tolerance. Four main findings emerged: first, we identify robust genetic correlations between suicide-related behaviors. Second, SUDs were correlated with individual suicide-related behaviors both in bivariate analyses and via a single SUD factor. Third, the genetic correlation between the SUD factor and suicide-related behaviors withstood adjustment for genetic contributions from depression and risk tolerance. Fourth, despite a large degree of similarity across correlations, some SUDs appeared to be more closely genetically related to certain suicide-related behaviors, indicating possible specificity. Similarly, depression showed variable degrees of association with the different suicide-related behaviors.

While genetic sharing amongst SUDs has been shown to be well captured by a common genetic factor, the genetics of specific suicide-related behaviors has remained unexplored. Our study finds support for a common genetic factor of suicide-related behaviors, although the model did not appear to represent the data well because the genetics of self-harm were indistinguishable from the genetics of suicide attempt; assessing the three suicide behaviors as separate correlates of the SUD factor ensured that the genetics of suicide death were also represented. While this high genetic overlap between self-harm and suicide attempt is consistent with twin studies (Richmond-Rakerd et al., 2019), this high r_g_ might also be attributable to “item contamination”, i.e., the self-harm GWAS utilized responses to an item broadly querying self-harming behaviors that included non-suicidal self-harm as well as suicide attempts. Indeed, of the 5,099 individuals endorsing the item, about half (N=2,658) reported a suicide attempt. Future studies that successfully parse suicidal ideation, persistent ideation and planning, suicide attempts, and non-suicidal self-injury might provide better insight into whether these behaviors, along with suicide death, represent a common genetic construct.

From the perspective of genetic overlap between the SUD factor and suicide, despite modeling individual correlations between the SUD factor and each suicide-related behavior, we were able to constrain these correlations to be equal. Thus, the specific genetic factors contributing to the correlations between SUDs and suicide-related behaviors may be distinct, while their effects are similar in magnitude. Despite this equality, the bivariate genetic correlations hinted at specific relationships between some SUDs and aspects of suicide-related behaviors. For example, the genetic correlation between OUD and suicide death (r_g_ = 0.45) appeared higher than the genetic correlations between the other SUDs and suicide death (r_g_s ranging from 0.33-0.40), although these correlations were not significantly different (likely due in part to the small sample size of the suicide death GWAS relative to the other GWAS). Although speculative, we hypothesize that this modestly higher genetic correlation might reflect the greater severity of OUD relative to the other SUDs included in our analysis, with OUD also potentially reflecting more severe comorbid polydrug misuse (as indexed by its high genetic correlation with all the other SUDs). There is substantial evidence to suggest that the number of substances used is more predictive of suicide than the specific type of substance used (Borges et al., 2000), and risk for suicide increases as the number of SUDs increases (Borges et al., 2000; Hakansson et al., 2011; Polimanti et al., 2021; G. W. Smith et al., 2011). It is estimated that of individuals with OUD, greater than 90% have used more than two other substances and over 25% met criteria for at least two comorbid SUDs within the same time period (Hassan & Le Foll, 2019). Another possibility is that some proportion of the suicide death cases were due to drug overdoses, which may also have contributed to the higher genetic correlation between suicide death and OUD (Docherty et al., 2020). Irrespective of the mechanism contributing to this nominally higher genetic correlation, our findings – while inconclusive – hint at the role of both shared and specific genetic relationships between individual SUDs and suicide-related behaviors.

The genetic correlations between SUDs and suicide-related behaviors remained partially independent of their individual genetic associations with both depression and risk tolerance. Unsurprisingly, the genetic correlations between depression and suicide-related behaviors (e.g., r_g_ = 0.66 for self-harm) were significantly stronger than the genetic correlations between depression and SUDs (e.g., r_g_ = 0.32 for CUD). Intriguingly, the associations between depression and the specific suicide-related behaviors significantly differed (i.e., applying equality constraints to these associations resulted in a worse-fitting model). Depression was most strongly associated with ever self-harmed and least associated with suicide death; this provides further evidence that the genetic contributions to suicide death may differ somewhat from the genetics of preceding stages like non-suicidal self-harm and attempt. Importantly, a weakness of existing GWAS of suicide attempt and death is that they lack sufficient power to probe intermediate phenotypes such as perseveration (persistent ideation and/or attempts), planning, intent, and history of attempts for individual suicide deaths (Witt et al., 2021). Alternatively, the genetic correlation between suicide death and depression may vary according to a number of factors (age, gender, comorbid psychiatric diagnoses, causes of suicide death), and the current finding may be specific to the genetic underpinnings of suicide death in Utah.

Given the plethora of research which describes risk-taking as a crucial component of SUDs (Dick et al., 2010; Hanson et al., 2014; Schneider et al., 2012; J. L. Smith et al., 2014), we expected to observe stronger genetic correlations between risk tolerance and SUDs compared to risk tolerance and suicide-related behaviors. However, risk tolerance appears to contribute similarly to both SUDs and suicide-related behaviors. For instance, the strongest genetic correlation between risk tolerance and an SUD (CUD; r_g_ = 0.40) could be statistically equated to the weakest genetic correlation between risk tolerance and a suicide-related behavior (ever self-harmed; r_g_ = 0.27; equality-p = 0.09). This finding re-affirms the role of risk tolerance in genetic predisposition to suicide-related behaviors. However, risk tolerance was broadly defined in the respective GWAS and likely represents positive urgency (i.e., disinhibition in the context of pleasure – the typical understanding of “takes risks”; Cyders et al., 2014). In contrast, many self-harming behaviors have been linked to negative urgency, or the propensity to act impusively in the context of negative affect (e.g., *When I am upset, I often act without thinking*; Anestis & Joiner, 2011). Large GWAS of negative urgency are currently lacking; future studies that include measures of risk in the context of negative urgency may identify stronger associations with suicide-related behaviors than we did in the current study.

We note several limitations: first, while we find evidence of genetic overlap between SUDs and suicide-related behaviors, we did not determine an underlying biological mechanism or identify causal relationships amongst these traits. Recent Mendelian randomization analyses propose a causal role of substance use on suicide attempt (Orri et al., 2020); however, in the current study we did not find evidence of causality, possibly due to no true association, or some of our phenotypes may have been underpowered to detect an association. Future analyses with larger samples will be better positioned to continue to investigate potential causal relationships. Additionally, significant gender differences have been documented between males and females for suicide-related behaviors, but we did not explore any sex differences in the current analysis. While women are more likely to attempt suicide (Nock et al., 2008) and men are more likely to die by suicide (Beautrais, 2001), it is not yet known whether genetic influences underlying these traits may differ by sex. Furthermore, SUD samples including data from the MVP (PAU and OUD) may unintentionally reflect gender differences as well, as most MVP participants are male. As more genetic data for SUDs and suicide-related behaviors becomes available, the genetic contributions to these traits should be examined in sex-stratified analyses. Similarly, as samples become more diverse, genetic analyses of these traits should expand beyond samples of European ancestry. Finally, because of differences in how the SUD phenotypes were assessed, with CUD and OUD being clinical diagnoses (DSM or ICD codes), PAU including a short screener for alcohol problems (the AUDIT-P), and FTND representing a score from a questionnaire assessing nicotine dependence, these phenotypes may represent different ranges of severity, with CUD and OUD only representing the most severe, diagnosed cases.

Overall, the current study extends previous research by demonstrating genetic covariance between four SUDs, three suicide-related behaviors, depression and risk tolerance. These findings raise the possibility of future efforts aimed at identifying shared genetic pathways of risk. At present, they underscore the importance of the transdiagnostic role of suicide-related behaviors. Indeed, the genetic correlation between some SUDs and suicide-related behaviors exceeded the genetic correlations between those of SUDs and depression, a comorbidity that is frequently evaluated in clinical settings. Our study confirms that individuals with genetic liability for SUDs may also be at higher risk for suicide-related behaviors.

## Data Availability

All publicly available GWAS summary statistics used in the analyses can be accessed via their original publications. For access to the suicide death summary statistics from the Utah Suicide Genetics Research Study, please contact Dr. Docherty.

## Acknowledgements

This work was supported by grant YIG-0-064-18 from the American Foundation for Suicide Prevention. The authors thank all investigators and participants who have contributed to the publicly available GWAS data used in this study. The authors thank Million Veteran Program (MVP) staff, researchers, and volunteers, who have contributed to MVP, and especially participants who previously served their country in the military and now generously agreed to enroll in the study. (See https://www.research.va.gov/mvp/ for more details). The citation for MVP is Gaziano, J.M. et al. Million Veteran Program: A mega-biobank to study genetic influences on health and disease. J Clin Epidemiol 70, 214-23 (2016). This research is based on data from the Million Veteran Program, Office of Research and Development, Veterans Health Administration, and was supported by the Veterans Administration (VA) Cooperative Studies Program (CSP) award #G002.

## Footnotes

1 The LDSC estimator is unbounded and can produce r_g_ estimates outside of –1 to 1 due to sampling variation. See https://groups.google.com/g/ldsc_users/c/3jtyM4mmTGs

