## Supplemental Figures for "Exploring the genetic overlap of suicide-related behaviors and substance use disorders"

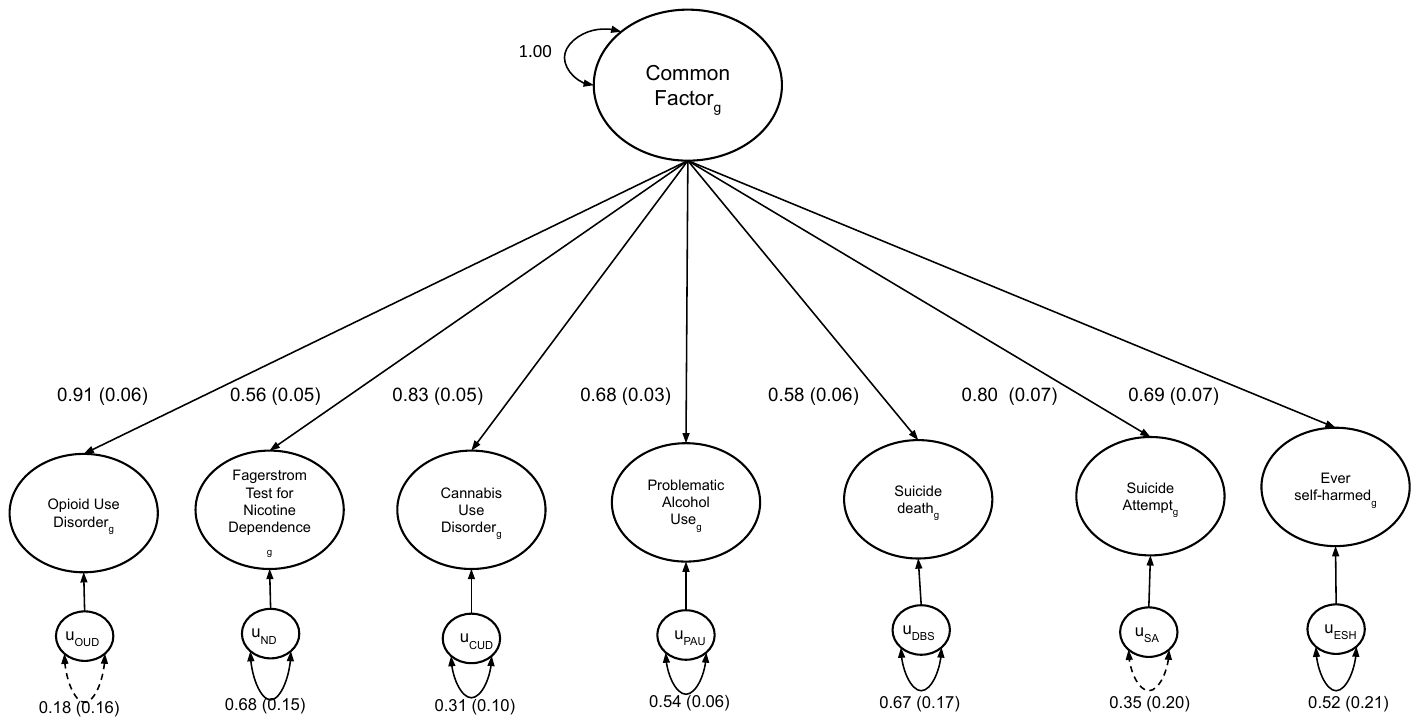

**Figure S1. Common factor model results.** Parameter estimates are standardized with standard errors in parentheses. Solid black lines are used to indicate significance (p < 0.05) while dotted black lines represent non-significant paths. Model fit was adequate (CFI > 0.90 and SRMR < 0.15): χ^2^(14) = 33.99, p_chisq_ = 0.002, AIC = 61.99, CFI = 0.967, SRMR = 0.119.

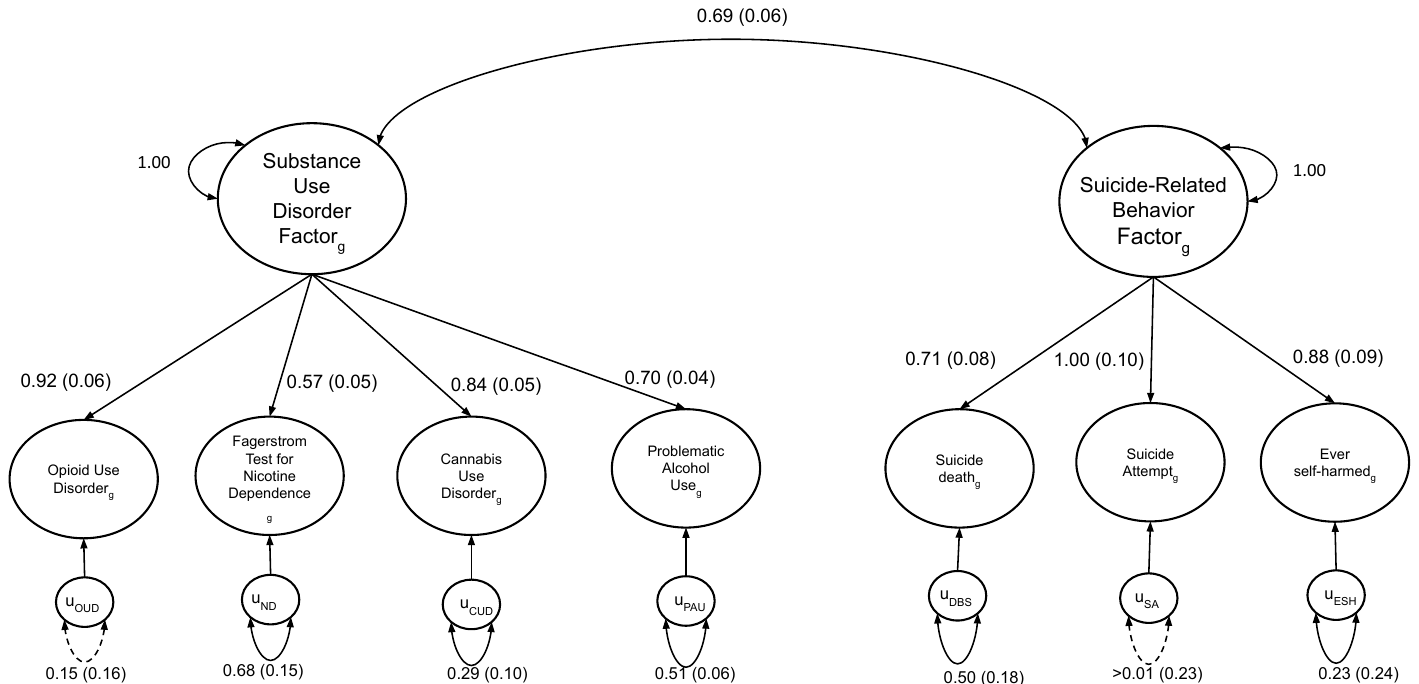

**Figure S2. Correlated two-factor model results.** Parameter estimates are standardized with standard errors in parentheses. Solid black lines are used to indicate significance (p < 0.05) while dotted black lines represent non-significant paths. Model fit was good (CFI > 0.95 and SRMR < 0.08): χ^2^(13) = 13.90, p_chisq_ = 0.38, AIC = 43.90, CFI = 0.99, SRMR = 0.065.

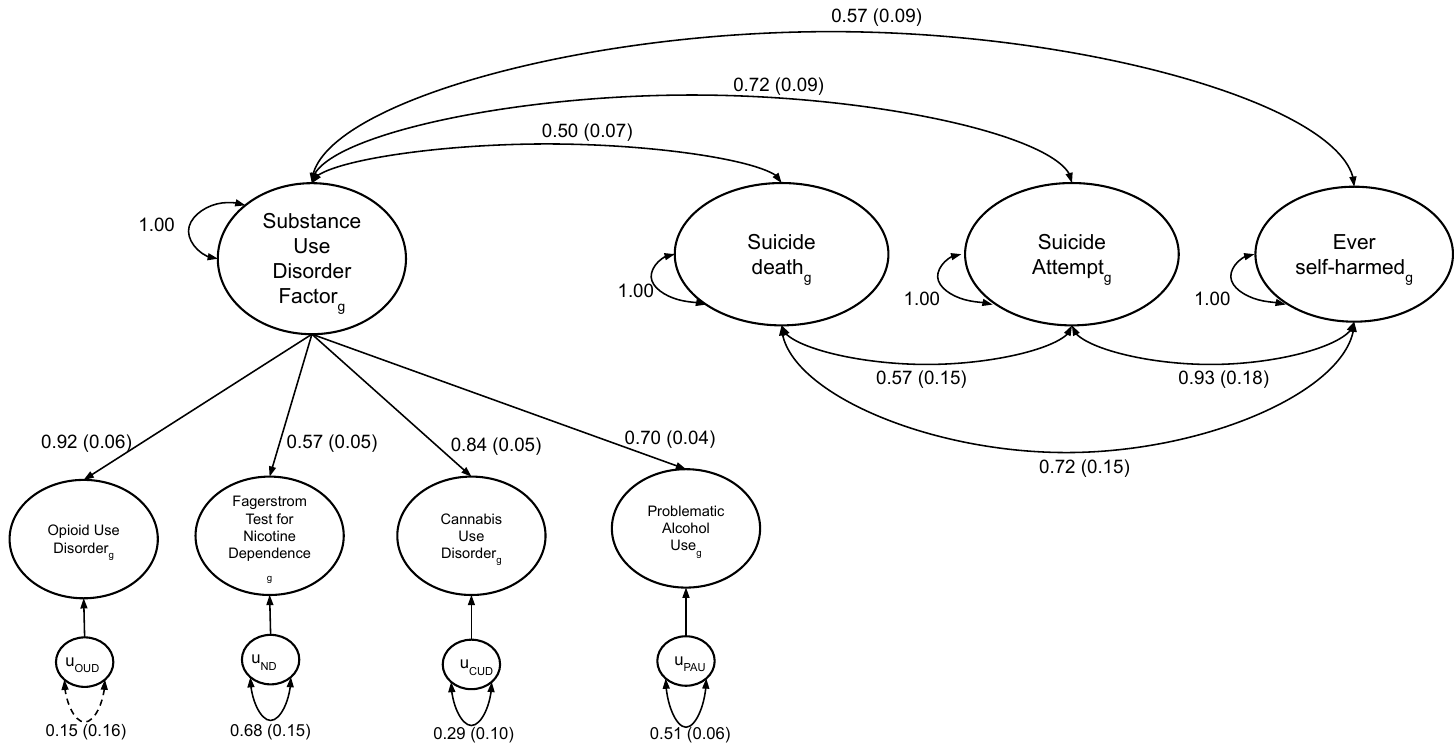

**Figure S3. Common SUD and specific suicide-related behaviors model results.** Parameter estimates are standardized with standard errors in parentheses. Solid black lines are used to indicate significance (p < 0.05) while dotted black lines represent non-significant paths. Model fit was good (CFI > 0.95 and SRMR < 0.08): χ^2^(11) = 10.06, p_chisq_ = 0.53, AIC = 44.06, CFI = 1, SRMR = 0.053.

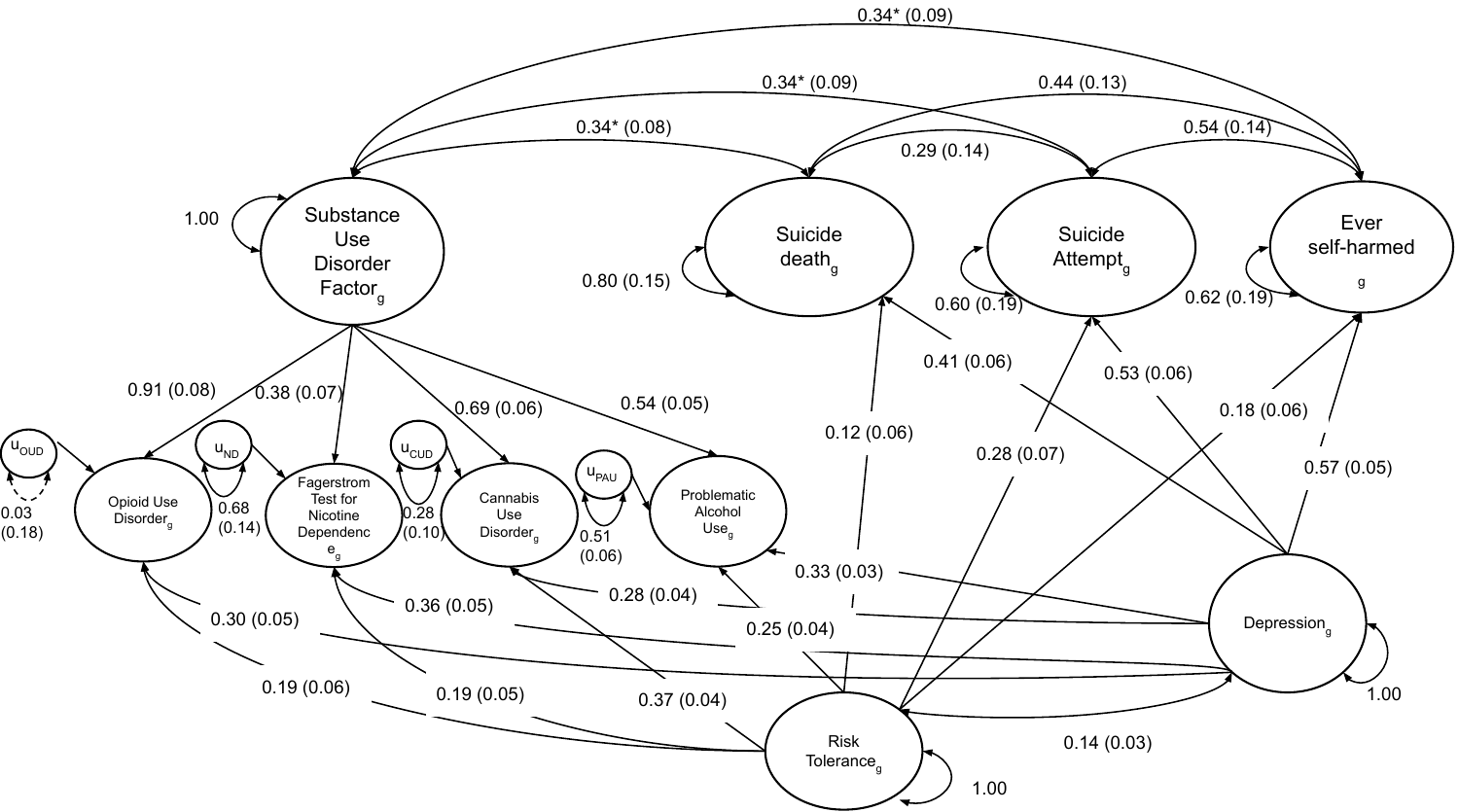

**Figure S4. Common SUD and specific suicide-related behaviors model with covariates and equality constraints on the correlations between SUD factor and suicide-related behaviors.** Correlations between the SUD factor and all three suicide-related behaviors were constrained to be equal, indicated by an asterisk (*). Parameter estimates are standardized with standard errors in parentheses. Solid black lines are used to indicate significance (p < 0.05) while dotted black lines represent non-significant paths. Model fit was good (CFI > 0.95 and SRMR < 0.08): χ^2^(13) = 13.43, p_chisq_ = 0.41, AIC = 77.43, CFI = 0.998, SRMR = 0.042.

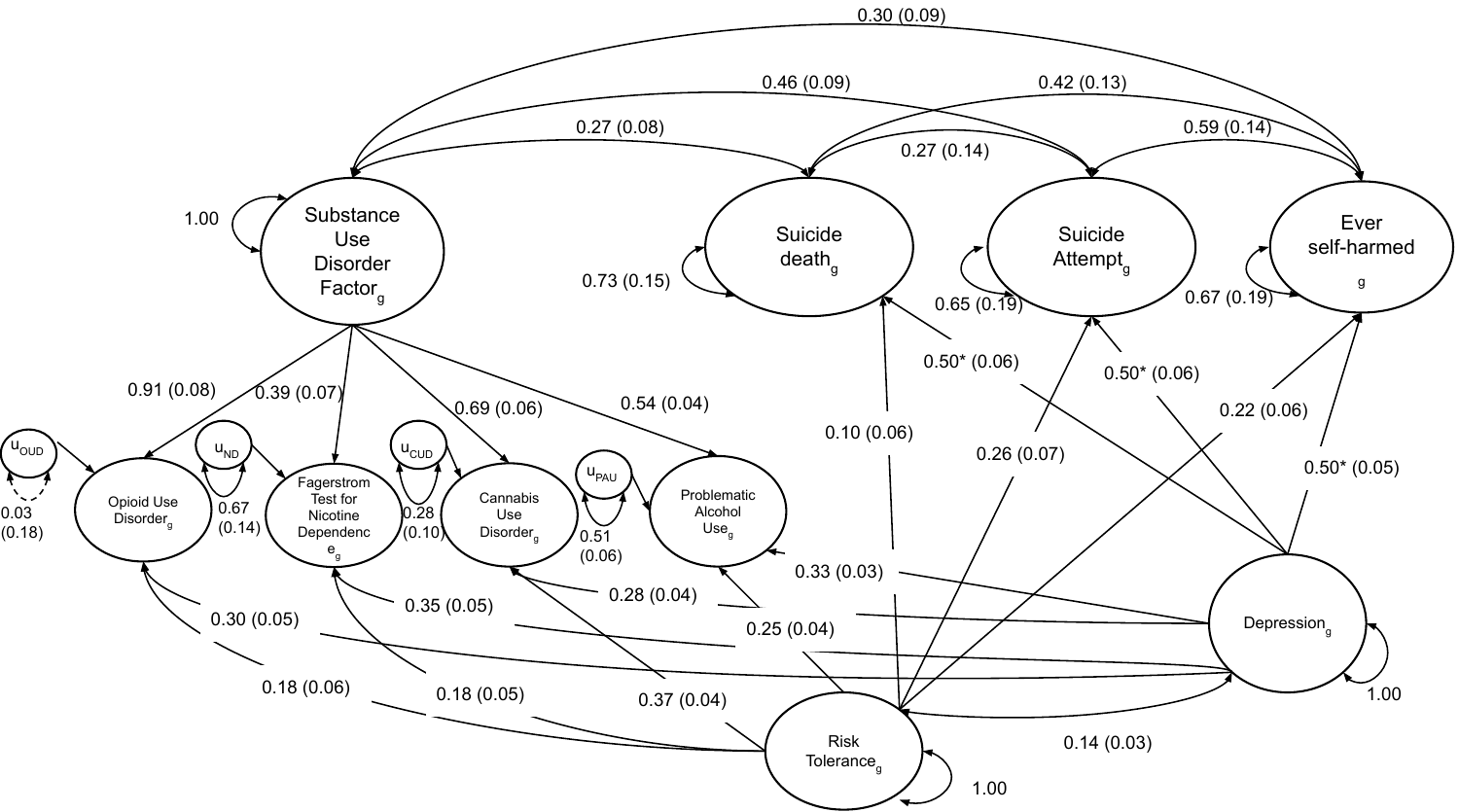

**Figure S5. Common SUD and specific suicide-related behaviors model with covariates and equality constraints on the associations between depression and suicide-related behaviors.** Associations between depression and all three suicide-related behaviors were constrained to be equal, indicated by an asterisk (*). Parameter estimates are standardized with standard errors in parentheses. Solid black lines are used to indicate significance (p < 0.05) while dotted black lines represent non-significant paths. Model fit was good (CFI > 0.95 and SRMR < 0.08): χ^2^(13) = 22.44, p_chisq_ = 0.05, AIC = 86.44, CFI = 0.991, SRMR = 0.043.

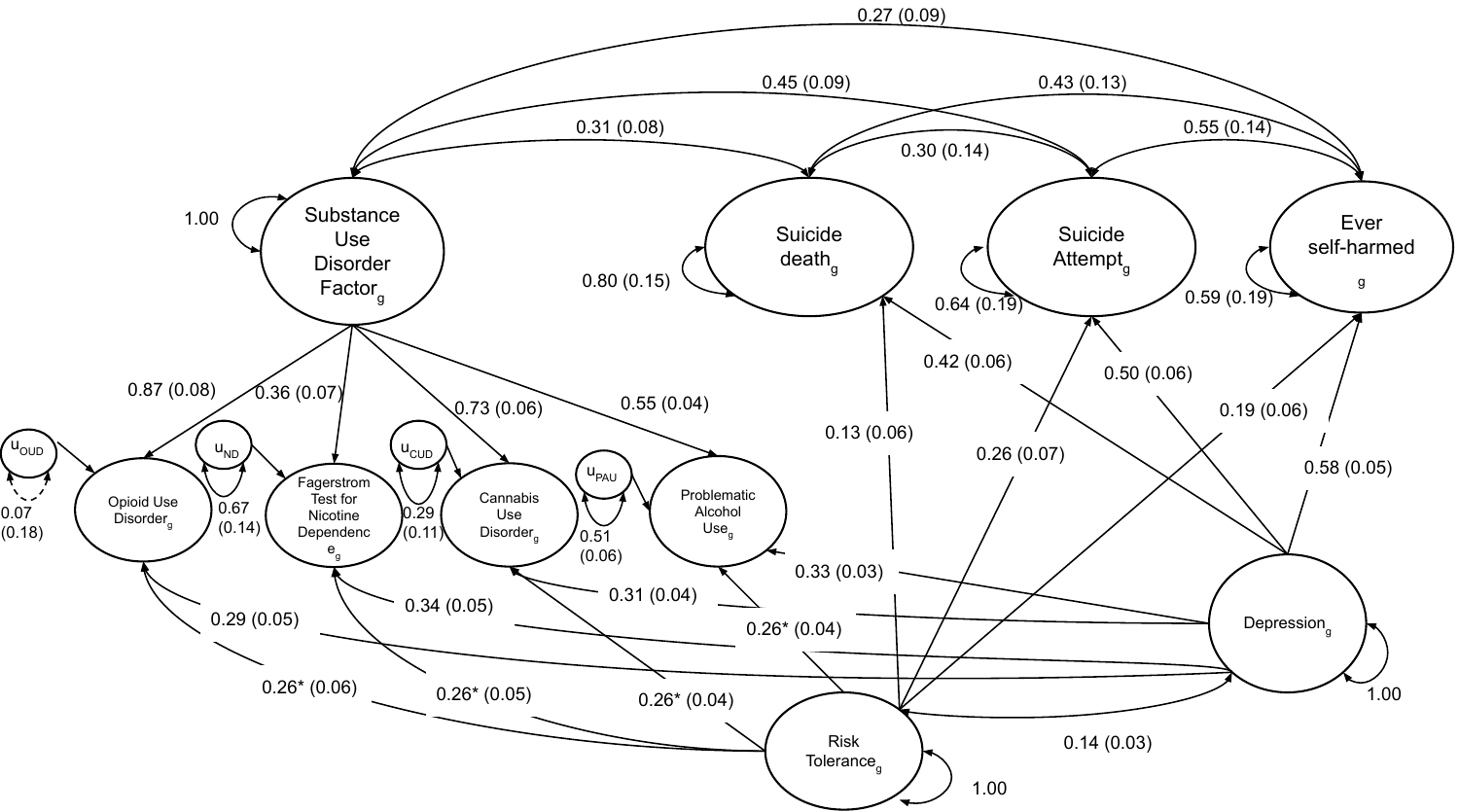

**Figure S6. Common SUD and specific suicide-related behaviors model with covariates and equality constraints on the associations between risk tolerance and SUDs.** Associations between risk tolerance and all four SUDs were constrained to be equal, indicated by an asterisk (*). Parameter estimates are standardized with standard errors in parentheses. Solid black lines are used to indicate significance (p < 0.05) while dotted black lines represent non-significant paths. Model fit was good (CFI > 0.95 and SRMR < 0.08): χ^2^(14) = 49.08, p_chisq_ = 8.71e^-6^, AIC = 111.08, CFI = 0.969, SRMR = 0.047.

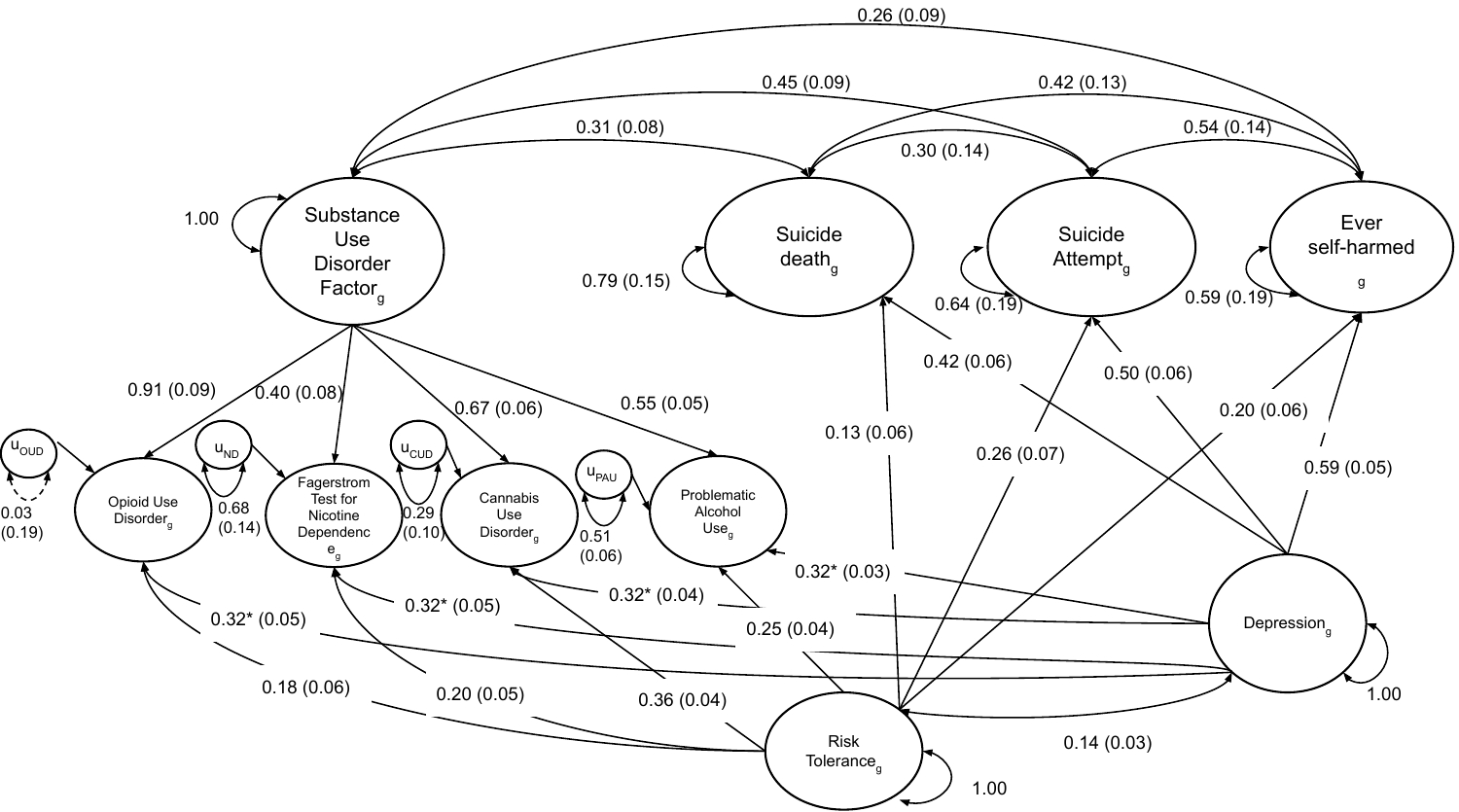
 **Figure S7. Common SUD and specific suicide-related behaviors model with covariates and equality constraints on the associations between depression and SUDs.** Associations between depression and all four SUDs were constrained to be equal, indicated by an asterisk (*). Parameter estimates are standardized with standard errors in parentheses. Solid black lines are used to indicate significance (p < 0.05) while dotted black lines represent non-significant paths. Model fit was good (CFI > 0.95 and SRMR < 0.08): χ^2^(14) = 18.49, p_chisq_ = 0.19, AIC = 80.49, CFI = 0.994, SRMR = 0.042.

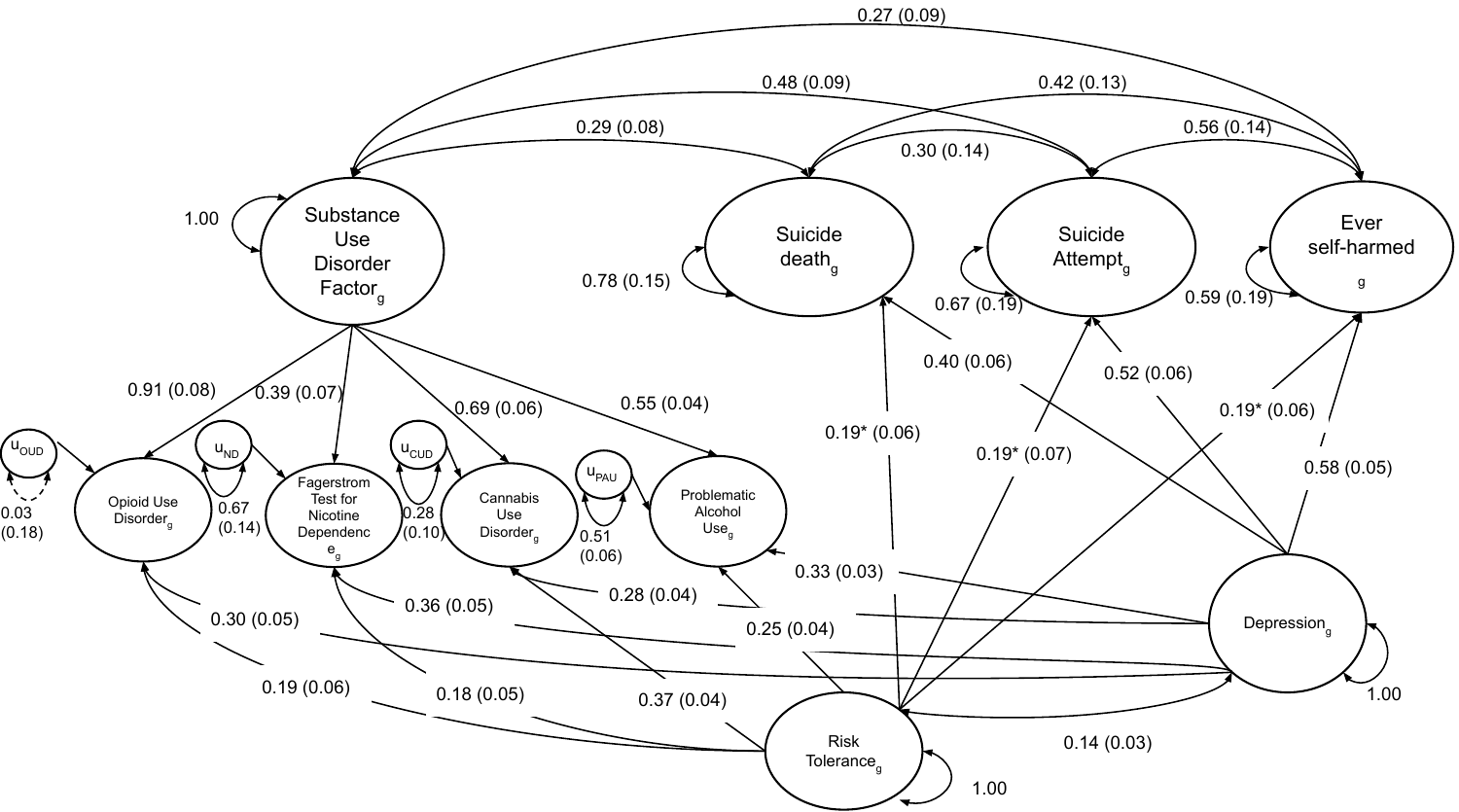

**Figure S8. Common SUD and specific suicide-related behaviors model with covariates and equality constraints on the associations between risk tolerance and suicide-related behaviors.** Associations between risk tolerance and all three suicide-related behaviors were constrained to be equal, indicated by an asterisk (*). Parameter estimates are standardized with standard errors in parentheses. Solid black lines are used to indicate significance (p < 0.05) while dotted black lines represent non-significant paths. Model fit was excellent (CFI > 0.99 and SRMR < 0.04): χ^2^(13) = 14.33, p_chisq_ = 0.35, AIC = 78.33, CFI = 0.997, SRMR = 0.039.
